# Widespread Self-Medication and Unsafe Access to Analgesics and NSAIDs in Urban Conakry, Guinea: Structural Determinants, Epidemiological Patterns, and Health System Implications – A Cross-Sectional Study of 1,032 Participants

**DOI:** 10.64898/2026.05.21.26353180

**Authors:** Daouda Lawa Garandji, Alpha Oumar Balde

## Abstract

**Background:** Self-medication with analgesics and non-steroidal anti-inflammatory drugs (NSAIDs) is common in low- and middle-income countries and may expose users to preventable adverse outcomes. Evidence from Guinea remains scarce. This study aimed to estimate the prevalence of self-medication with analgesics and NSAIDs among pharmacy clients in urban Conakry, identify associated factors, and describe clinical risk situations.

**Methods:** We conducted a pharmacy-based analytical cross-sectional study in 30 private pharmacies across Conakry, Guinea. A total of 1,032 participants seeking analgesics or NSAIDs were enrolled between November 3, 2012, and April 5, 2013. Self-medication was defined as acquisition or use without a valid medical prescription. Factors associated with self-medication were analysed using multivariable logistic regression.

**Results:** Among 1,032 participants, 603 reported self-medication (prevalence 58.4%). Previous unsupervised use was reported by 78.7%. The most frequently used medicines were paracetamol (56.9%, n=587), diclofenac (21.3%, n=220), ibuprofen (17.9%, n=185), and aspirin (3.9%, n=40). Overall, 68.0% (n=702) reported no knowledge of potential adverse effects. Clinical risk situations were frequent: gastrointestinal disorders (41.3%, n=426), hypertension (9.2%, n=95), and pregnancy exposure among reproductive-age women (26.0%). In multivariable analysis, self-medication was independently associated with previous analgesic/NSAID use (aOR = 2.8, 95% CI: 2.1–3.6), lack of knowledge of adverse effects (aOR = 1.9, 95% CI: 1.4–2.5), informal occupation (aOR = 1.6, 95% CI: 1.2–2.2), and age 18–59 years (aOR = 1.5, 95% CI: 1.1–2.1).

**Conclusions:** In this pharmacy-based study conducted in urban Conakry, self-medication with analgesics and NSAIDs was common and frequently associated with limited awareness of potential adverse effects. These findings support the need for strengthened pharmaceutical regulation, pharmacist-led counselling, health literacy interventions, and improved access to primary care.

## I. INTRODUCTION

Self-medication, commonly defined as the use of medicines without medical prescription or professional clinical assessment, is an important public health issue in low- and middle-income countries (LMICs) [1-4]. It may improve access to treatment for minor ailments and reduce pressure on health services, but it also exposes users to misdiagnosis, inappropriate dosing, prolonged use, drug interactions, adverse drug reactions, and delayed access to appropriate care [2-4]. These risks are particularly relevant for commonly used medicines such as analgesics and non-steroidal anti-inflammatory drugs (NSAIDs), which are often perceived as harmless despite well-established safety concerns [5-9].

Analgesics and NSAIDs are among the most frequently used medicines for fever, headache, musculoskeletal pain, joint pain, and influenza-like symptoms [5-9]. Their wide availability, relatively low cost, previous user experience, and frequent over-the-counter access contribute to their widespread use. However, unsupervised use may lead to excessive dosing, repeated intake, inappropriate combinations, and use despite contraindications [5].

Paracetamol is generally safe when used at recommended doses, but overdose or repeated supratherapeutic dosing can cause severe hepatotoxicity and acute liver failure [14-18]. NSAIDs, including diclofenac, ibuprofen, and aspirin, are associated with gastrointestinal bleeding, peptic ulcer complications, renal injury, increased blood pressure, and cardiovascular events [19-24]. These risks are particularly important among individuals with gastrointestinal disorders, hypertension, renal disease, older age, or pregnancy exposure [36-38].

In sub-Saharan Africa, self-medication is shaped not only by individual behaviour but also by structural determinants, including limited access to affordable primary care, weak pharmaceutical regulation, informal medicine markets, high consultation costs, and variable access to pharmacist counselling [10-13]. Studies from Ethiopia, Eritrea, Tanzania, Ghana, Burkina Faso, and other African settings suggest that self-medication is common and often associated with limited knowledge of medicine-related risks [10-12].

In Guinea, published pharmaco-epidemiological evidence on analgesic and NSAID self-medication remains scarce. We therefore conducted a pharmacy-based cross-sectional study in urban Conakry to estimate the prevalence of self-medication with analgesics and NSAIDs, identify associated factors, and describe clinical risk situations. The study was conducted in 2012–2013 and is presented as historical baseline evidence. Its interpretation focuses on structural determinants of self-medication, including access barriers, low risk awareness, and weak regulation of medicine access, rather than on estimating current prevalence.

## II. METHODS

### Study design and setting

We performed a cross-sectional analytical study following the STROBE guidelines. The study took place in Conakry, the capital of Guinea, a densely populated urban area with a pluralistic pharmaceutical system comprising regulated private pharmacies, semi-regulated outlets, and informal markets. Data collection occurred in 30 private pharmacies distributed across the main communes between November 3, 2012, and April 5, 2013.

### Participants

We included individuals aged ≥15 years who were requesting or purchasing at least one analgesic or NSAID, able to provide verbal informed consent, and capable of responding to the questionnaire. We excluded those purchasing on behalf of another person, refusing to participate, or unable to respond due to cognitive impairment or severe illness.

### Sample size and sampling

We enrolled 1,032 participants using consecutive sampling: all eligible individuals presenting to the participating pharmacies during the study period were invited. This approach minimises selection bias in high-flow outpatient pharmacy settings.

### Data collection

Trained pharmacy staff and research assistants administered a standardised, pre-tested structured questionnaire developed from WHO guidelines on rational drug use and previous pharmacoepidemiological studies. The questionnaire captured sociodemographic characteristics, drug utilisation patterns, indications, comorbidities, knowledge of adverse effects, and source of recommendation.

### Variable definitions (STROBE-compliant)

- Primary outcome: self-medication (binary) – acquisition or use of an analgesic or NSAID without a valid medical prescription.
- Secondary outcomes: type of drug used; knowledge of adverse effects (yes/no); high-risk use – defined as use despite a known contraindication or comorbidity (gastrointestinal disease, hypertension, pregnancy) or lack of knowledge of adverse effects.
- Exposure variables: age, sex, occupation, education, prior use of analgesics, source of recommendation, comorbidities.

### Bias control

Selection bias was reduced by consecutive inclusion across multiple pharmacies. Information bias was minimised through a standardised questionnaire and trained interviewers. Recall bias was limited by focusing on current or recent medication use. Confounding was addressed in multivariable regression.

### Statistical analysis

Analyses were performed using Stata/R. Descriptive statistics (frequencies, percentages, means ± SD) described drug utilisation and self-medication prevalence. Bivariate associations used chi-square tests for categorical variables and Student’s t-test or Mann-Whitney U test for continuous variables. Variables with p < 0.20 entered a multivariable logistic regression model with self-medication as the dependent variable. Results are presented as adjusted odds ratios (aOR) with 95% confidence intervals (CI). Model validation included assessment of multicollinearity (variance inflation factor), goodness-of-fit (Hosmer-Lemeshow test), and discrimination (area under the ROC curve). A sensitivity analysis used high-risk NSAID use as the outcome.

### Ethical considerations

This study was conducted in accordance with the ethical principles of the Declaration of Helsinki. Ethical approval for the study protocol was granted by the National Ethics Committee for Health Research of the Republic of Guinea (Comité National d’Éthique pour la Recherche en Santé — Authorization No 007/CNERS/12.

Institutional administrative authorization for the doctoral research was also obtained from the Department of Pharmaceutical Sciences, Faculty of Medicine, Pharmacy and Odontostomatology, Gamal Abdel Nasser University of Conakry, Guinea (Authorization No. 033/2012). Doctoral thesis defense was authorized by Rectoral Order No. 446/RECT/2013. The doctoral thesis was publicly defended on November 20, 2013, at the Faculty of Medicine, Pharmacy and Odontostomatology, Chair of Pharmacology, Clinical Pharmacy and Therapeutics, Gamal Abdel Nasser University of Conakry-Guinea.

Before participation, all individuals were informed of the study’s objectives and procedures. The ethics committee approved the use of verbal informed consent due to the anonymous, low-risk nature of the cross-sectional survey, as documented in the study protocol. Verbal informed consent was obtained from each participant and documented prior to inclusion. Participation was voluntary, anonymous, and confidential. No personally identifiable information was collected.

## III. RESULTS

### Participant characteristics

We included 1,032 participants. The population was predominantly male (55.4%, n = 572) and aged 18–59 years (45.5%). Informal workers (traders, labourers) represented 39.9% (n = 412), students 25.0% (n = 258) (Table 1).

**Table 1.** Sociodemographic characteristics of study participants (N = 1,032)

| Variable | n | % |
| --- | --- | --- |
| Total participants | 1,032 | 100.0 |
| Male | 572 | 55.4 |
| Female | 460 | 44.6 |
| Age < 18 years | 455 | 44.1 |
| Age 18–59 years | 470 | 45.5 |
| Age ≥ 60 years | 107 | 10.3 |
| Informal workers | 412 | 39.9 |
| Students | 258 | 25.0 |
| Other occupations | 362 | 35.1 |

### Prevalence of self-medication

Overall, 58.4% (603/1,032) reported self-medication. Importantly, 78.7% (812/1,032) had previously used analgesics or NSAIDs without a prescription, indicating a recurrent pattern. Self-medication was more frequent among adults aged 18–59, informal workers, and those with prior drug exposure (Table 2).

**Table 2.**
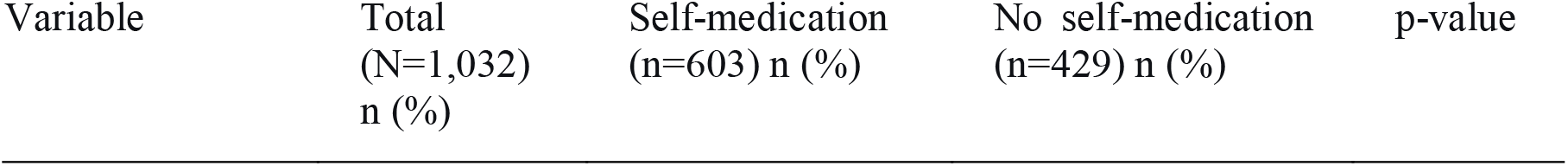
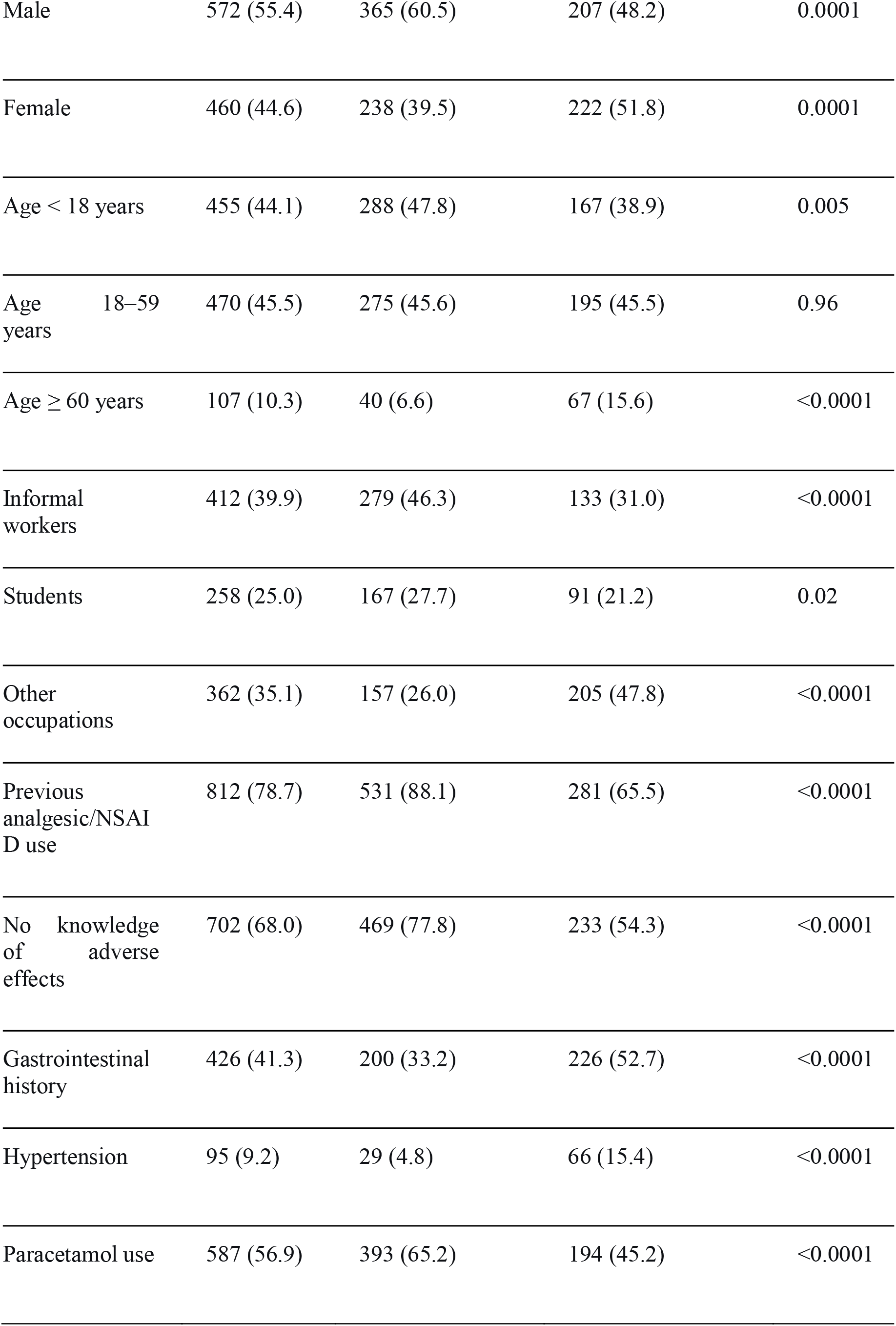
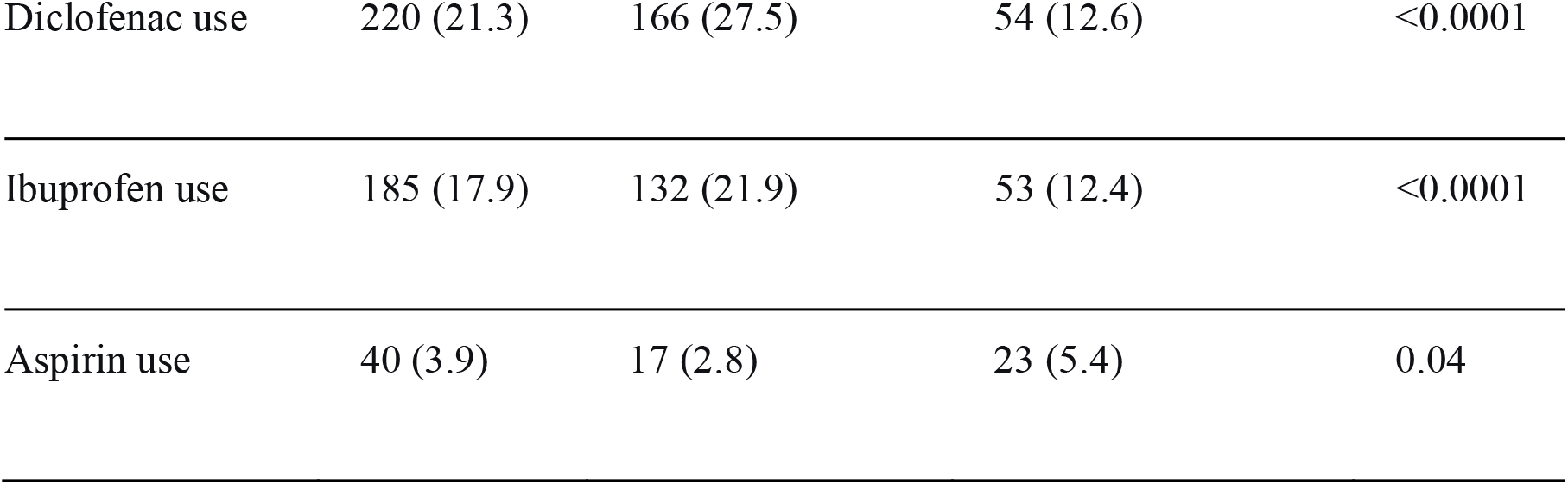
Characteristics of participants according to self-medication status.

### Drug utilisation patterns

Paracetamol was the most commonly used drug (56.9%, n=587), followed by diclofenac (21.3%, n=220), ibuprofen (17.9%, n=185), and aspirin (3.9%, n=40) (Table 3). Diclofenac and ibuprofen were disproportionately represented in the self-medication group.

**Table 3.** Drug utilisation patterns among study participants.

| Medicine | n | % |
| --- | --- | --- |
| Paracetamol | 587 | 56.9 |
| Diclofenac | 220 | 21.3 |
| Ibuprofen | 185 | 17.9 |
| Aspirin | 40 | 3.9 |
| Total | 1,032 | 100.0 |

### Indications

The main indications were fever/flu-like symptoms (19.6%, n=202), headache (17.0%, n=175), joint pain (15.7%, n=162), and low back pain (9.3%, n=96), suggesting predominantly symptomatic use (Table 4).

**Table 4.** Reported clinical indications for analgesic or NSAID use.

| Indication | n | % |
| --- | --- | --- |
| Fever/influenza-like symptoms | 202 | 19.6 |
| Headache | 175 | 17.0 |
| Joint pain | 162 | 15.7 |
| Low back pain | 96 | 9.3 |
| Other indications | 397 | 38.5 |
| Total | 1,032 | 100.0 |

### Knowledge and risk awareness

A total of 68.0% (702/1,032) were unaware of potential adverse effects of analgesics or NSAIDs. Lack of awareness was significantly associated with lower education, self-medication, and informal employment (Table 5).

**Table 5.** Risk awareness and clinical risk situations.

| Variable | n | % |
| --- | --- | --- |
| No knowledge of potential adverse effects | 702 | 68.0 |
| History of gastrointestinal disorder | 426 | 41.3 |
| Hypertension | 95 | 9.2 |
| Pregnancy exposure among reproductive-age women* | 68 | 26.0 |
| Clinical high-risk use (e.g., CKD, liver disease, bleeding risk) | 124 | 12.0 |

\*Percentage calculated among women of reproductive age (n=262 based on female self-medication group approximation).

### Determinants of self-medication

Multivariable logistic regression identified four independent predictors: prior analgesic use (aOR = 2.8, 95% CI: 2.1–3.6), lack of knowledge of adverse effects (aOR = 1.9, 95% CI: 1.4–2.5), informal occupation (aOR = 1.6, 95% CI: 1.2–2.2), and age 18–59 years (aOR = 1.5, 95% CI: 1.1–2.1). Higher education and physician consultation were protective (Table 6).

**Table 6.** Multivariable logistic regression analysis of factors associated with self-medication.

| Variable | Crude OR | 95% CI | p-value | Adjusted OR | 95% CI | p-value |
| --- | --- | --- | --- | --- | --- | --- |
| Previous analgesic/NSAID use | 3.9 | 2.9–5.2 | <0.001 | 2.8 | 2.1–3.6 | <0.001 |
| No knowledge of adverse effects | 2.9 | 2.2–3.8 | <0.001 | 1.9 | 1.4–2.5 | <0.001 |
| Informal occupation | 1.9 | 1.5–2.5 | <0.001 | 1.6 | 1.2–2.2 | 0.002 |
| Age 18–59 years | 1.6 | 1.2–2.1 | 0.001 | 1.5 | 1.1–2.1 | 0.010 |
| Male sex | 1.7 | 1.3–2.1 | <0.001 | 1.4 | 1.1–1.9 | 0.015 |
| Education level<br>(≤ primary) | 1.3 | 1.0–1.7 | 0.045 | 1.2 | 0.9–1.6 | 0.120 |
| Physician<br>consultation<br>(past 6 months) | 0.6 | 0.5–0.8 | <0.001 | 0.7 | 0.5–0.9 | 0.008 |
| Gastrointestinal<br>history | 0.4 | 0.3–0.5 | <0.001 | 0.5 | 0.4–0.7 | <0.001 |
| Hypertension | 0.3 | 0.2–0.5 | <0.001 | 0.4 | 0.2–0.6 | <0.001 |

Model diagnostics: Hosmer-Lemeshow test: $\chi^2 = 8.23$ , $df = 8$ , $p = 0.41$ ; Area under ROC curve = 0.74; Maximum VIF = 1.32.

## IV. FIGURES

**Figure 1.**
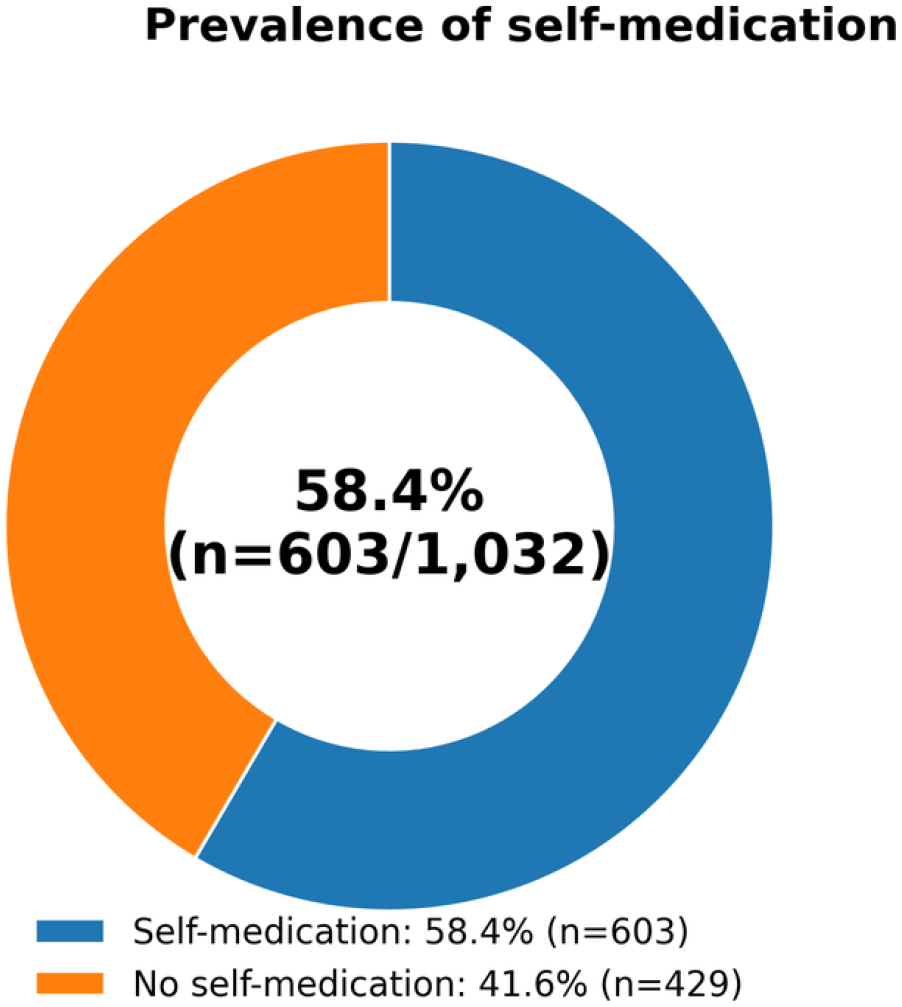
Prevalence of self-medication Proportion of participants reporting self-medication (58.4%, n=603/1,032).

**Figure 2.**
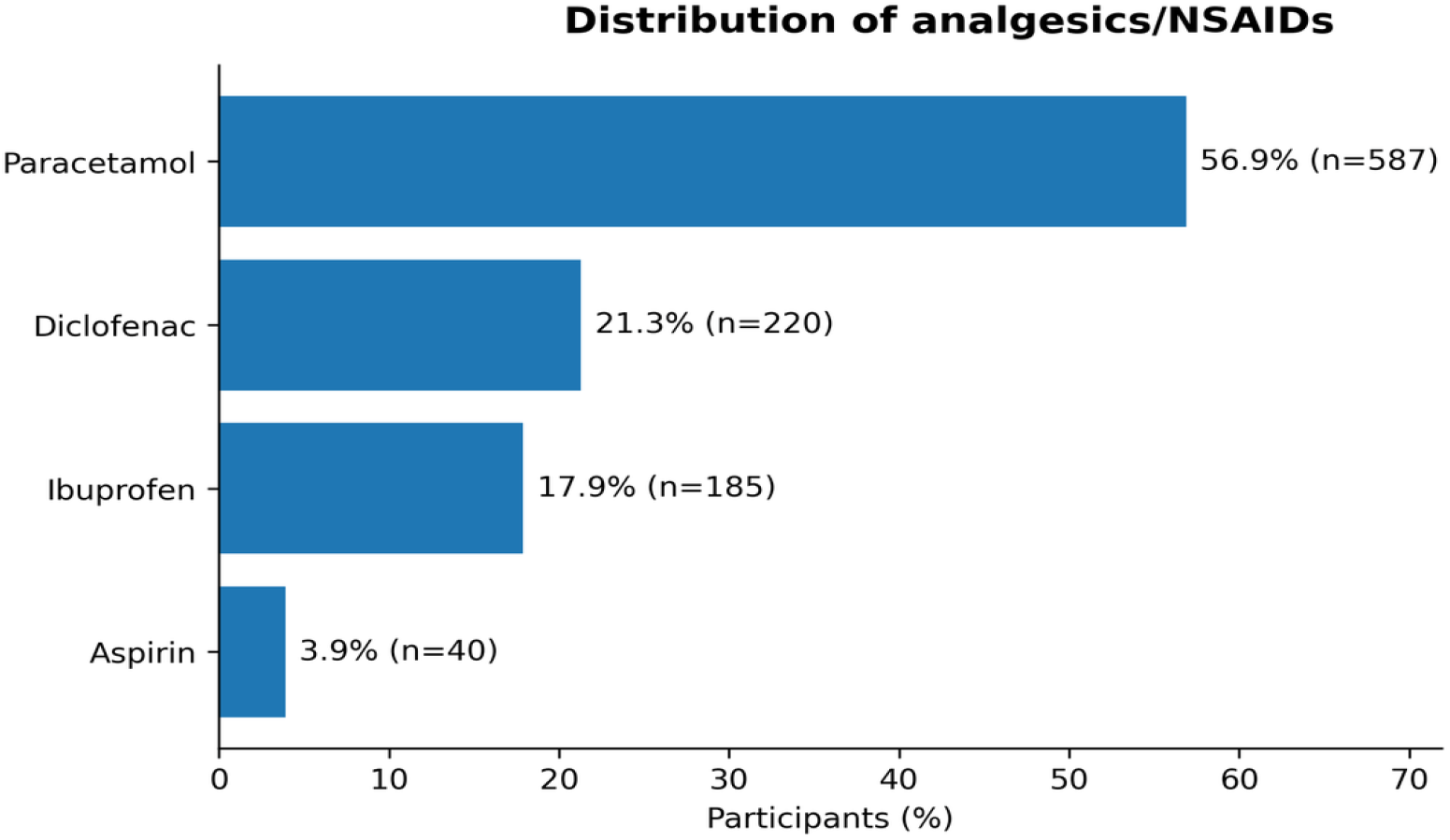
Distribution of analgesics/NSAIDs Paracetamol (56.9%, n=587), diclofenac (21.3%, n=220), ibuprofen (17.9%, n=185), aspirin (3.9%, n=40).

**Figure 3.**
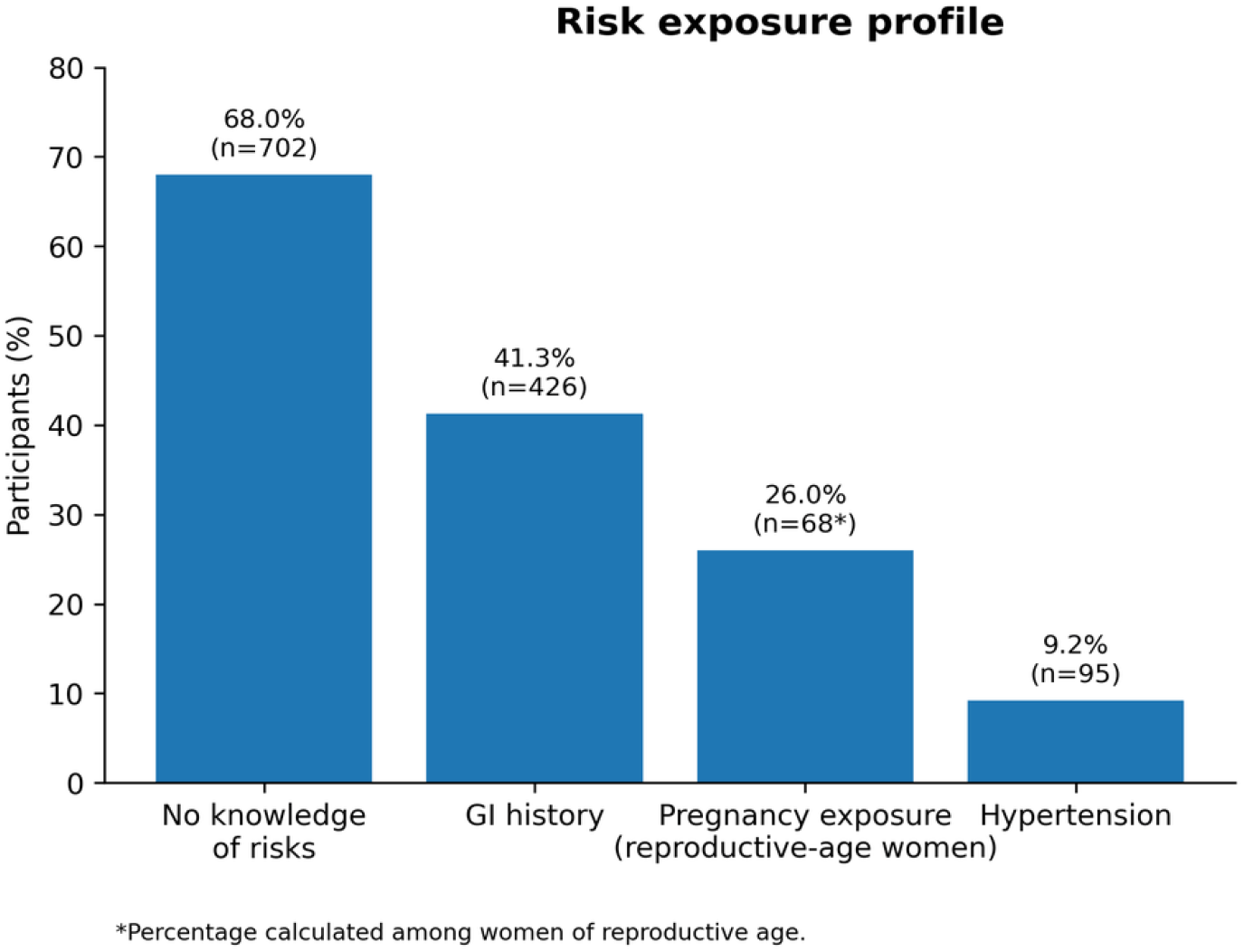
Risk exposure profile GI history (41.3%, n=426), hypertension (9.2%, n=95), pregnancy exposure (26.0% of reproductive-age women), no knowledge of risks (68.0%, n=702).

**Figure 4.**
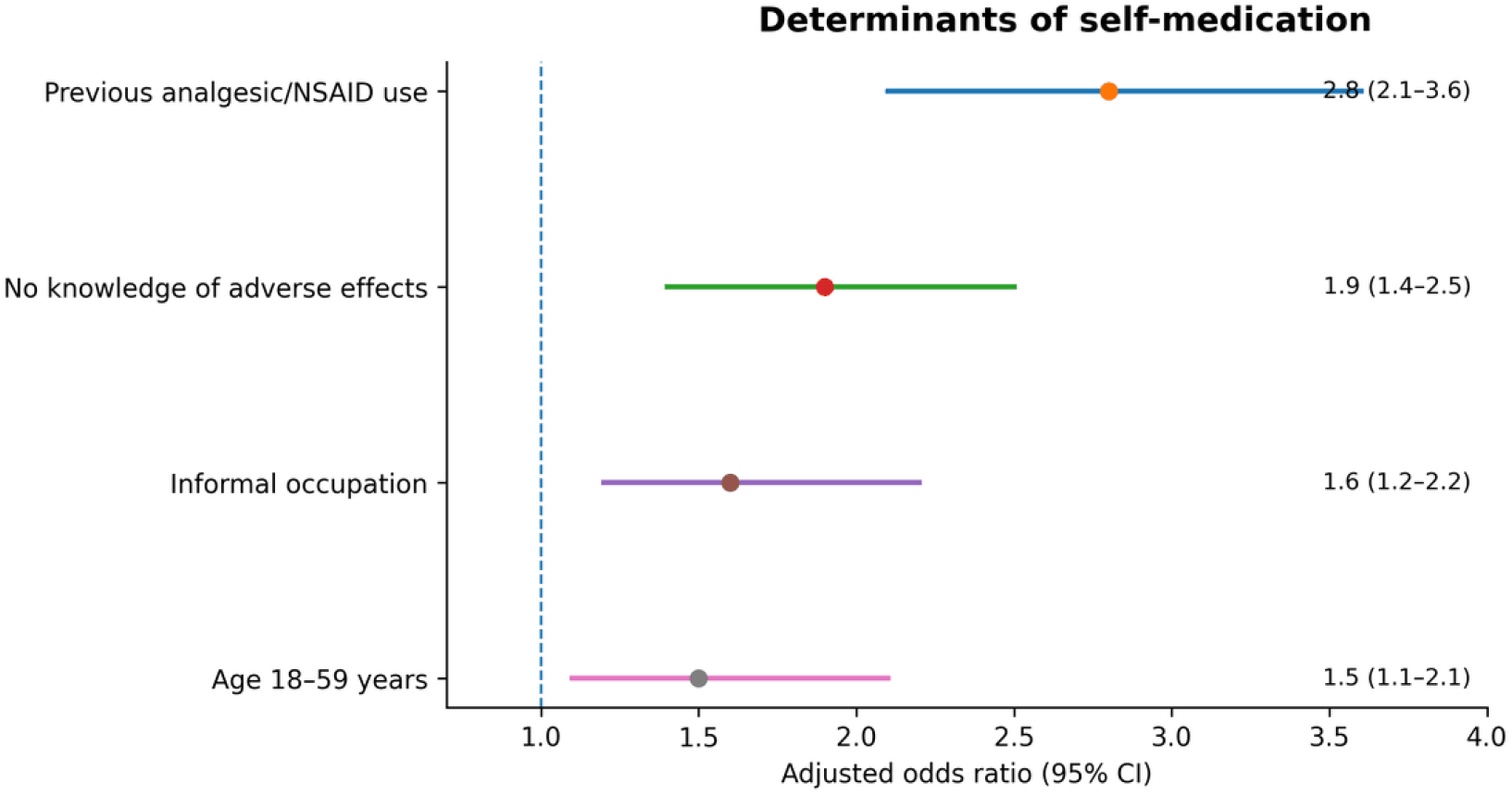
Determinants of self-medication Forest plot of adjusted odds ratios from Table 6 (previous use, no knowledge, informal job, age 18–59).

**Figure 5.**
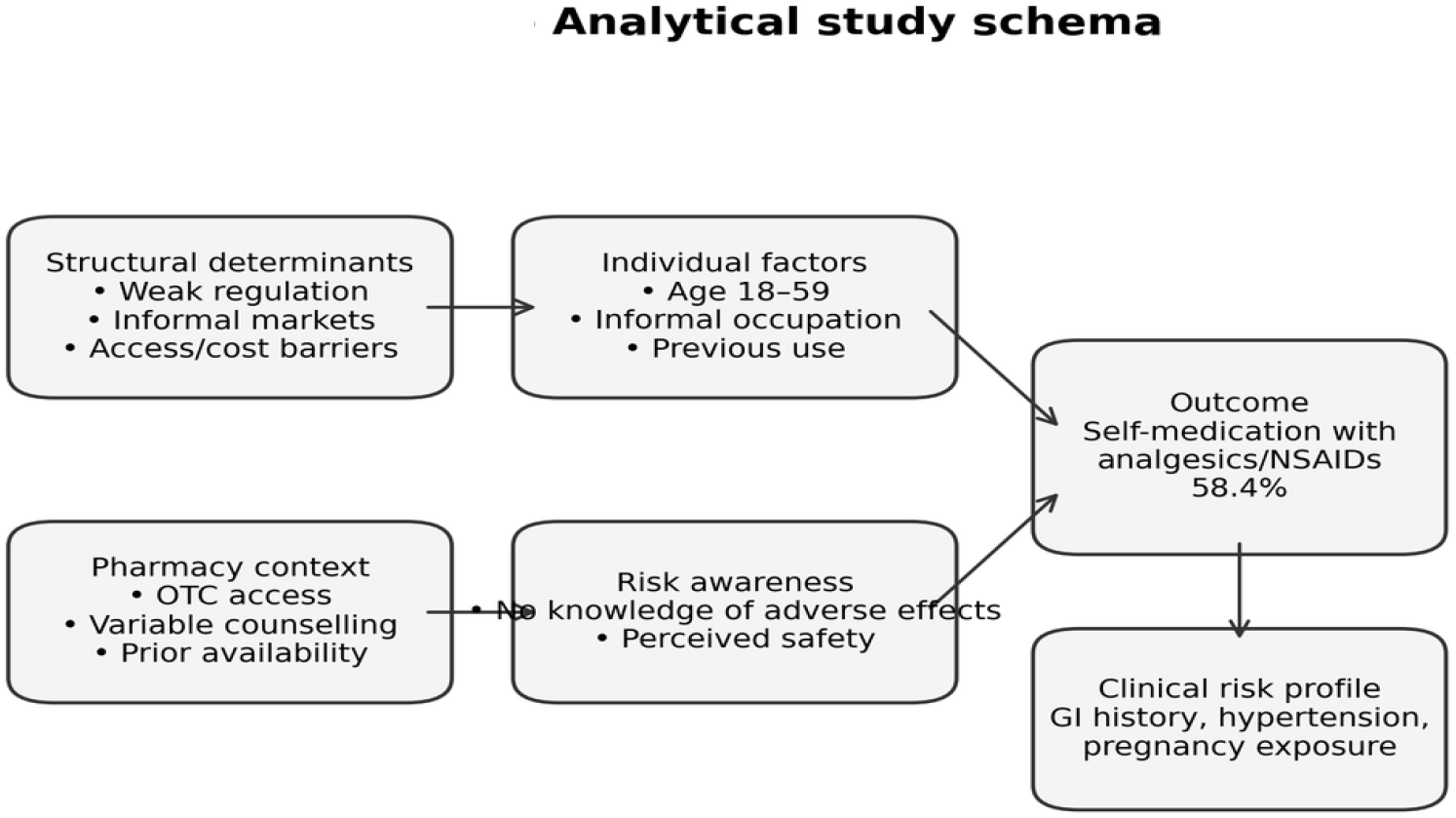
Analytical study schema Conceptual framework showing structural determinants and individual factors associated with self-medication.

**Figure 6.**
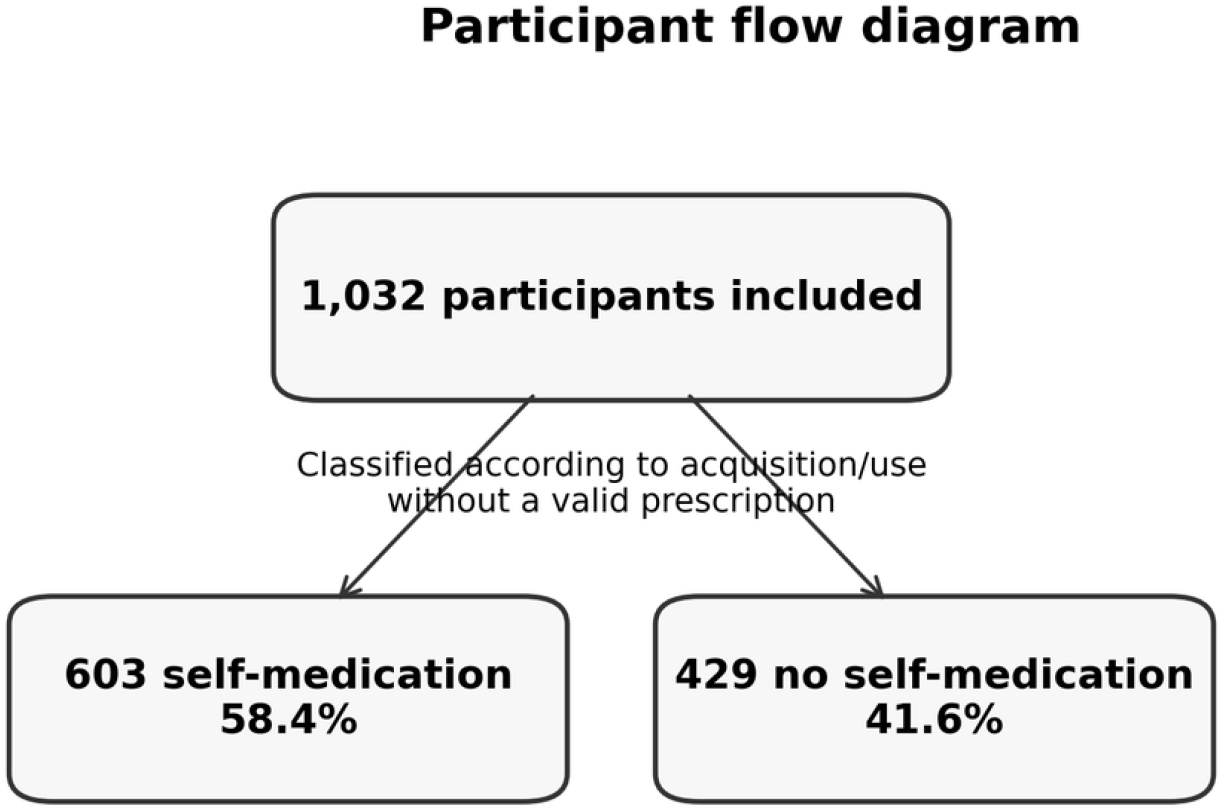
Participant flow diagram 1,032 included → 603 self-medication → 429 no self-medication.

## V. DISCUSSION

### Principal findings

In this large cross-sectional study of urban Conakry, self-medication with analgesics and NSAIDs was highly prevalent (58.4%), recurrent (78.7% prior use), and often unsafe. High-risk use was common among individuals with gastrointestinal disorders (41.3%), hypertension (9.2%), and pregnant women (26.0% of reproductive-age women). Only one-third of participants were aware of potential adverse effects (32.0%). These findings point to a systemic failure in pharmaceutical governance, health literacy, and primary care access.

### Comparison with other settings

The observed prevalence is at the upper range of LMIC estimates (55–80%) [1-4], similar to reports from urban Nigeria and Ethiopia [10-12]. The predominance of paracetamol and NSAIDs mirrors global patterns [5-9], but the high frequency of diclofenac (21.3%) in self-medication is concerning given its cardiovascular and gastrointestinal risks [19-21]. A global overview of NSAID consumption confirms wide regional variations, with over□the□counter availability driving higher use in LMICs [39].

### Clinical risk burden

The finding that 41.3% of individuals with gastrointestinal disease use NSAIDs is alarming given the well-established association with upper gastrointestinal bleeding [19-21]. Similarly, use among hypertensive individuals (9.2%) disregards cardiovascular risks [24-28]. The 26.0% exposure during pregnancy is particularly worrisome given links to miscarriage and fetal harm [36-38]. Paracetamol-related hepatotoxicity and NSAID-related renal injury are also likely under-recognised in this setting [14-18,29-32]. Beyond paracetamol, NSAIDs themselves can cause idiosyncratic drug□induced liver injury, ranging from mild transaminase elevation to acute liver failure, although the absolute risk is lower than for paracetamol [33-35].

### Structural determinants

Our findings confirm that self-medication is driven not by individual choice alone but by weak regulation, easy access to pharmacies and informal markets, and low health literacy. Pharmacies, despite being a key interface, miss opportunities for counselling – only a minority of self-medicating participants received pharmacist guidance. Moreover, the circulation of substandard and falsified medicines further exacerbates safety concerns [13]. A systematic review of medicine quality in low□income settings found that poor storage conditions, lack of regulatory enforcement, and fragmented supply chains frequently lead to substandard analgesics, further compounding the risks of self□medication [40].

### Strengths and limitations

The study included a large sample size and multiple pharmacy sites, allowing assessment of real-world medicine use patterns in an under-documented setting.

Several limitations should be considered. First, the cross-sectional design precludes causal interpretation. Second, consecutive sampling and pharmacy-based recruitment may introduce selection bias and limit external validity. Third, residual clustering at pharmacy level may persist because dispensing practices may vary across pharmacies. Fourth, the study was conducted in 2012–2013, which may limit direct applicability to current prevalence estimates. However, structural determinants of medicine access and regulation in Guinea have remained relatively stable, and the study is presented as historical baseline evidence. Finally, the study did not directly assess pharmacist counselling quality, medicine storage conditions, or exposure to informal drug markets outside pharmacies.

### Implications for policy and practice

Based on these results, we recommend: (1) enforce prescription requirements for high-risk NSAIDs; (2) regulate informal drug markets; (3) launch public awareness campaigns targeting low-literacy populations; (4) integrate mandatory pharmacist counselling for over-the-counter analgesics; (5) establish community-based pharmacovigilance. These actions align with WHO rational use of medicines goals.

## Conclusion

Self-medication with analgesics and NSAIDs in urban Conakry is widespread, structurally embedded, and frequently unsafe. Multi-level interventions targeting regulation, education, and primary care are urgently needed.

## Data Availability

The datasets generated and/or analysed during the current study are not publicly available due to ethical restrictions (participants' anonymity and confidentiality) but are available from the corresponding author on reasonable request.

## Authors’ contributions

Daouda Lawa GARANDJI conceptualised and designed the study, supervised data collection, performed the statistical analysis, and drafted the initial manuscript. Alpha Oumar BALDE critically revised the manuscript for important intellectual content. All authors read and approved the final manuscript and agree to be accountable for all aspects of the work.

## REFERENCES

1. Hughes CM, McElnay JC, Fleming GF. Benefits and risks of self-medication. Drug Saf. 2001;24(14):1027–37.

2. Cooper RJ. Over-the-counter medicine abuse: a review. J Subst Use. 2013;18(2):82–107.

3. Shehnaz SI, Agarwal AK, Khan N. Self-medication among adolescents. J Adolesc Health. 2014;55(4):467–83.

4. Ruiz ME. Risks of self-medication practices. Curr Drug Saf. 2010;5(4):315–23.

5. Koffeman AR, Valkhoff VE, Celik S, et al. High-risk use of non-steroidal anti-inflammatory drugs. Br J Gen Pract. 2014;64(621):e191–8.

6. Bianco A, Licata F, Zucco R, et al. Self-medication practices. Front Pharmacol. 2020;11:584069.

7. Montastruc F, Bondon-Guitton E, Abadie D, et al. Pharmacovigilance and self-medication. Therapie. 2016;71(2):257–62.

8. Perrot S, Citte J, Louis P, et al. Self-medication with over-the-counter analgesics. Eur J Pain. 2019;23(2):326–38.

9. Mehuys E, Gevaert P, Brusselle G, et al. Self-medication with over-the-counter analgesics: a survey. J Pain. 2019;20(2):215–23.

10. Ocan M, Obuku EA, Bwanga F, et al. Household antimicrobial self-medication. BMC Public Health. 2015;15:742.

11. Wafula FN, Goodman CA. Are interventions for improving the quality of drug shop services effective? Int J Qual Health Care. 2010;22(4):316–23.

12. Wafula FN, Miriti EM, Goodman CA. Examining characteristics, knowledge and regulatory practices of drug shops. BMC Health Serv Res. 2012;12:223.

13. Ozawa S, Evans DR, Bessias S, et al. Prevalence and estimated economic burden of substandard and falsified medicines. JAMA Netw Open. 2018;1(4):e181662.

14. Dart RC, Bailey E. Does therapeutic use of acetaminophen cause acute liver failure? Pharmacotherapy. 2007;27(9):1219–30.

15. Larson AM, Polson J, Fontana RJ, et al. Acetaminophen-induced acute liver failure. Hepatology. 2005;42(6):1364–72.

16. Nourjah P, Ahmad SR, Karwoski C, Willy M. Estimates of acetaminophen overdoses. Pharmacoepidemiol Drug Saf. 2006;15(6):398–405.

17. Lee WM. Acetaminophen toxicity. Hepatology. 2004;40(1):6–9.

18. Yoon E, Babar A, Choudhary M, et al. Acetaminophen-induced hepatotoxicity. J Clin Transl Hepatol. 2016;4(2):131–42.

19. Hernández-Díaz S, Rodríguez LA. Association between NSAIDs and upper gastrointestinal bleeding. Arch Intern Med. 2000;160(14):2093–9.

20. Lanas A, García-Rodríguez LA, Arroyo MT, et al. Risk of upper gastrointestinal ulcer bleeding. Gut. 2006;55(12):1731–8.

21. Massó González EL, Patrignani P, Tacconelli S, et al. Variability among NSAIDs and risk of GI bleeding. Arthritis Rheum. 2010;62(6):1592–601.

22. Bhatt DL, Scheiman J, Abraham NS, et al. ACCF/ACG/AHA guideline for NSAID GI risk. Circulation. 2008;118(18):1894–909.

23. Lanas A, Chan FK. Peptic ulcer disease. Lancet. 2017;390(10094):613–24.

24. Coxib and traditional NSAID Trialists’ (CNT) Collaboration. Vascular and upper gastrointestinal effects of NSAIDs. Lancet. 2013;382(9894):769–79.

25. Bally M, Dendukuri N, Rich B, et al. Risk of acute myocardial infarction with NSAIDs. BMJ. 2017;357:j1909.

26. Schmidt M, Christiansen CF, Mehnert F, et al. Non-aspirin NSAIDs and atrial fibrillation. BMJ. 2011;343:d3450.

27. Pope JE, Anderson JJ, Felson DT. Meta-analysis of NSAIDs and blood pressure. Arch Intern Med. 1993;153(4):477–84.

28. García Rodríguez LA, Tacconelli S, Patrignani P. Role of dose and duration of NSAID use. J Am Coll Cardiol. 2008;52(20):1628–36.

29. Ungprasert P, Cheungpasitporn W, Crowson CS, Matteson EL. Individual NSAIDs and risk of AKI. Eur J Intern Med. 2015;26(4):285–91.

30. Zhang X, Donnan PT, Bell S, Guthrie B. NSAIDs and risk of AKI: meta-analysis. BMC Nephrol. 2017;18:256.

31. Schneider V, Lévesque LE, Zhang B, et al. NSAIDs and risk of renal failure. Am J Epidemiol. 2006;164(9):881–9.

32. Whelton A. Nephrotoxicity of NSAIDs. Am J Med. 1999;106(5B):13S–24S.

33. Schmeltzer PA, Kosinski AS, Kleiner DE, et al. Liver injury from NSAIDs. Liver Int. 2016;36(4):603–9.

34. Andrade RJ, Lucena MI, Fernández MC, et al. Drug-induced liver injury. Gastroenterology. 2005;129(2):512–21.

35. Rubin R. Acetaminophen and liver failure. JAMA. 2019;321(4):327–9.

36. Kristensen DM, Hass U, Lesné L, et al. Analgesic use and reproductive disorders. Hum Reprod. 2011;26(1):235–44.

37. Rebordosa C, Kogevinas M, Horváth-Puhó E, et al. Acetaminophen use during pregnancy. Int J Epidemiol. 2009;38(3):706–14.

38. Nakhai-Pour HR, Broy P, Sheehy O, et al. NSAID use during pregnancy and miscarriage risk. CMAJ. 2011;183(15):1713–20.

39. McGettigan P, Henry D. Use of NSAIDs worldwide. PLoS Med. 2013;10(2):e1001388.

40. Almuzaini T, Choonara I, Sammons H. Substandard medicines review. BMJ Open. 2013;3:e002923.

